# A systematic review and meta-analysis of Long COVID symptoms

**DOI:** 10.1101/2022.03.08.22272091

**Authors:** Arun Natarajan, Ashish Shetty, Gayathri Delanerolle, Yutian Zeng, Yingzhe Zhang, Vanessa Raymont, Shanaya Rathod, Sam Halabi, Kathryn Elliot, Peter Phiri, Jian Qing Shi

**Affiliations:** Hillingdon Hospital NHS Foundation Trust; University College London Hospitals NHS Foundation Trust; University College London; University of Oxford, Nuffield Department of Primary Health Care Sciences; Southern University of Science and Technology; West China Hospital of Sichuan University; University of Oxford, Department of Psychiatry; Southern Health NHS Foundation Trust; O’Neill Institute for National and Global Health Law, Georgetown University; University of Southampton, Department of Psychology; The Alan Turing Institute, London

**Keywords:** Long Covid, Pain, Neuropsychiatry, Neurology, Autonomic Dysfunction, Gastrointestinal

## Abstract

**Background:** Ongoing symptoms or the development of new symptoms following a SARS-CoV-2 diagnosis has caused a complex clinical problem known as “:Long COVID”: (LC). This has introduced further pressure on global healthcare systems as there appears to be a need for ongoing clinical management of these patients. LC personifies heterogeneous symptoms at varying frequencies. The most complex symptoms appear to be driven by the neurology and neuropsychiatry spheres.

**Methods:** A systematic protocol was developed, peer reviewed and published in PROSPERO. The systematic review included publications from the 1^st^ of December 2019-30^th^ June 2021 published in English. Multiple electronic databases were used. The dataset has been analysed using a random-effects model and a subgroup analysis based on geographical location. Prevalence and 95% confidence intervals (CIs) were established based on the data identified.

**Results:** Of the 302 studies, 49 met the inclusion criteria, although 36 studies were included in the meta-analysis. The 36 studies had a collective sample size of 11598 LC patients. 18 of the 36 studies were designed as cohorts and the remainder were cross-sectional. Symptoms of mental health, gastrointestinal, cardiopulmonary, neurological, and pain were reported.

**Conclusions:** The quality that differentiates this meta-analysis is that they are cohort and cross-sectional studies with follow-up. It is evident that there is limited knowledge available of LC and current clinical management strategies may be suboptimal as a result. Clinical practice improvements will require more comprehensive clinical research, enabling effective evidence-based approaches to better support patients.

**Funding:** None

## Introduction

Global experience with a rapidly evolving and advanced strain of the coronavirus have led to over a million deaths since January 2020. The first case of SARS-CoV-2 was reported in China around December 2019. Healthcare systems have been under immense pressure to support both SARS-CoV-2 patients and survivors who continue to demonstrate various symptomatologies which appear to impact the overall quality of life and wellbeing. A report from the Center for Disease Control and Prevention (CDC) in the United States reported that patients recovered from SARS-CoV-2 have continuous symptoms of shortness of breath, fatigue, brain fog, cough, chest pain, stomach pain and headache. Bin Cao and colleagues reported that these complications appear to last for at least 6 months thus far (1). Similarly, Carfi and colleagues reported 87.4% of the survivors suffered from a variety of symptoms at post-60 days since the original SARS-CoV-2 diagnosis (2).

As SARS-CoV-2 survivors continue to share their experiences, clinical researchers hypothesize the continuation of complex symptomatologies for a longer period of time than initially anticipated (3). As are a result, several independent authorities have developed Long COVID guidelines, although the consensus continues to change with the changing evidence base from data gathered from patients. Therefore, a universally accepted Long COVID definition is yet to be elaborated, although a general overview is available. One such important guideline set is from the National Institute for Health and Care Excellence (NICE), which stipulates “:*Long COVID’ (LC) is commonly used to describe symptoms that continue or develop after acute SARS-CoV-2 diagnosis post-4 weeks*”: (4). The current research landscape exploring LC is also limited due to the varying reports of symptoms identified in clinical datasets that demonstrates it to be a ‘*moving target’* and it is challenging clinical researchers to guide clinicians on the most optimal steps to pursue when managing the clinical care of these patients. The World Health Organization’s (WHO) Novel Coronavirus Pneumonia Emergency Response Epidemiology Team describes LC as *a complex course of illness*. Therefore, pandemic policymaking itself requires evidence-based clinical research along with patient-reported outcomes and clinician experiences to be reported in an effective manner to channel a more holistic approach to optimise long-term clinical management. A key component appears to be the difference in LC symptoms between men and women, as reported by Mathew et al., who demonstrate that these observations are vital to understand, and that at present this is based particularly on clinician experience with limited pathophysiological and aetiology (5).

In this study, we conducted a meta-analysis of peer-reviewed and published data using a systematic approach to better understand LC from a neurological and neuropsychiatry perspective.

## Methods

A systematic methodology was developed, peer reviewed, and published in PROSPERO (CRD42021235351). The primary aim of this systematic review was to determine the prevalence of LC symptomatologies pertaining to neuropsychiatry, neurology, and pain. The secondary aim was to determine any other infrequently reported symptoms that may influence neuropsychiatry and/or neurology and/or pain diagnosis following LC. The Preferred Reporting Items for Systematic Reviews and Meta-Analysis (PRISMA) was used to report this study.

### Search strategy

Multiple databases of Embase, Pubmed, Science Direct, and ProQuest were used with multiple MeSH terms such as *nervous system diseases, autonomic central nervous system diseases, autonomic diseases, autonomic nervous system disorders, disorders of the autonomic nervous system, autonomic nervous system diseases, peripheral autonomic nervous system diseases, autonomic peripheral nervous system diseases, parasympathetic nervous system diseases, sympathetic nervous system diseases, headaches, migraine, headache after mental exertion, exertional headache, tension headache, cluster headache, intra cranial hypertension, temporal headache, retro-orbital headache, cervicogenic headache, chronic pain, fibromyalgia, back pain, erythromelalgia, endometriosis, intercostal neuralgia, leg pain, neuropathic pain, chronic pelvic pain, sciatica, muscle fatigue, metal fatigue, cognition, apathy, sleep arousal, sleep deprivation, sleep initiation and maintenance, anxiety, depression emotional lability*.

All studies and surveys were included in the Preliminary R1 round. The reviews and metanalysis identified were scrutinized for references that can be included in our metanalysis. A final set was arrived at looking at the possible relevance of the studies comprising of 302 studies. This was analysed as per PRISMA diagram in Figure 1 in the Results section.

**Figure 1:**
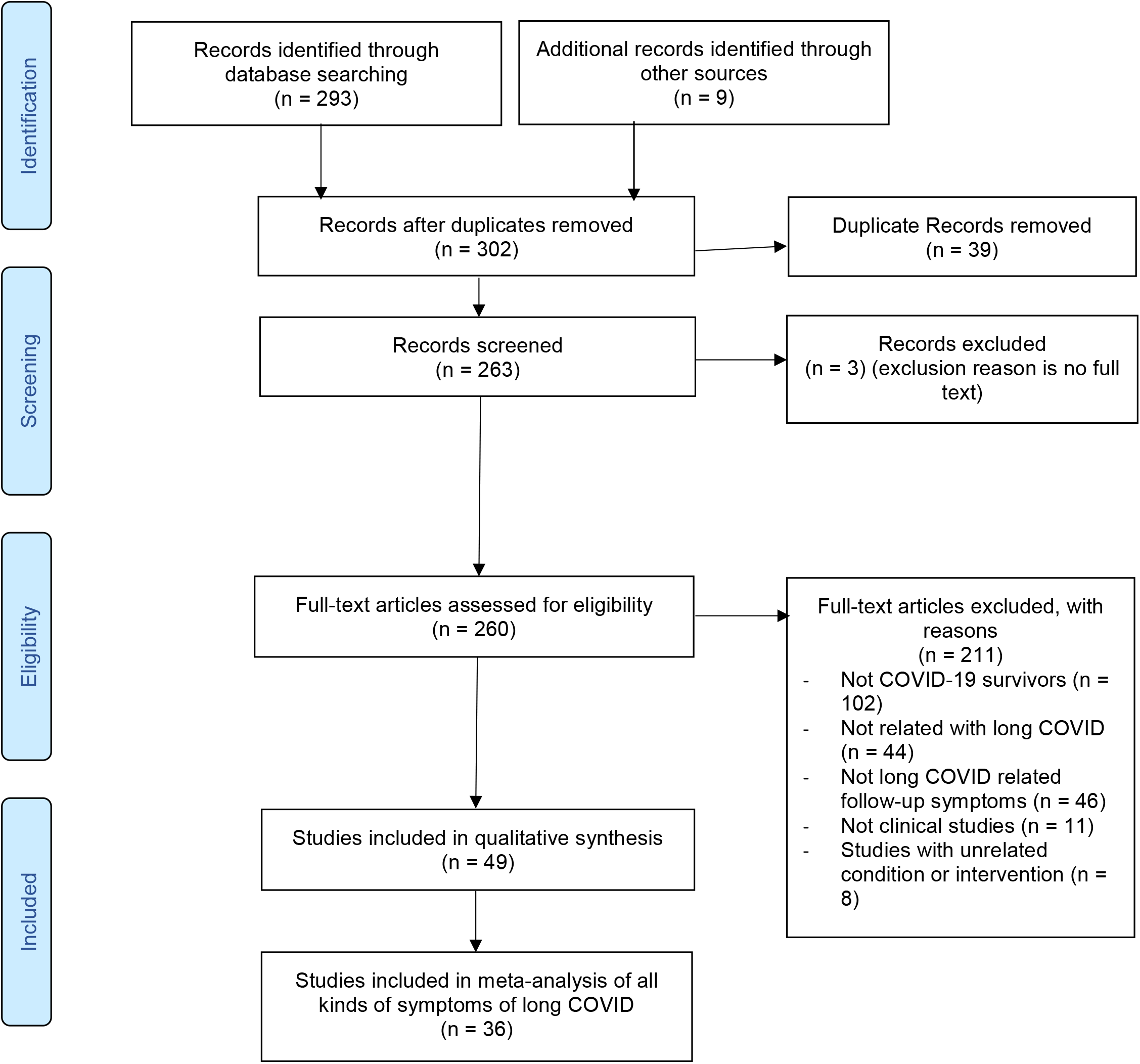
PRISMA Flow Diagram.

### Eligibility criteria

In this meta-analysis, we looked at persistent symptoms in COVID patients, including cohort and cross-sectional studies. All studies included were reported in English.

## Data extraction and synthesis

Screening and data extraction were performed by four independent reviewers. Any disagreements were discussed and reached a consensus by two reviewers. To fully investigate the impact of LC on the physical health of survivors, we grouped all reported symptoms into five main categories: general symptoms (which includes pain and other infrequently reported symptoms), neurological, mental disorders, cardiopulmonary, and obstetric problems.

Data extractions were made via studies that included SARS-CoV-2 survivors that had either been hospitalized or treated as outpatients. Therefore, these patients had a confirmed positive test for SARS-CoV-2 in addition to relevant symptoms. All studies that did not report on follow-up data were excluded. For studies that reported on a control and patient group, only the patient data was extracted and used. A data extraction sheet specific to the clinical question of this study was developed. This Excel spreadsheet included study type, sample size, country, characteristics, information, outcomes, duration of symptoms, and prevalence.

### Risk of bias assessment

A quality assessment was performed using the Newcastle-Ottawa Scale (NOS) (Table 1a) to critically appraise the literature included within the systematic review using common variables. Methodological quality and risk of bias was assessed by independent reviewers according to the NOS, which has validity for use in cohort studies (6) and the adapted version (7) for cross-sectional studies. The scale consists of eight items with three quality parameters: (i) selection, (ii) comparability, and (iii) outcome. We scored the quality of the studies (poor, fair, and good) by allocating stars to each domain as stated below:

**Table 1a [line 216]:**
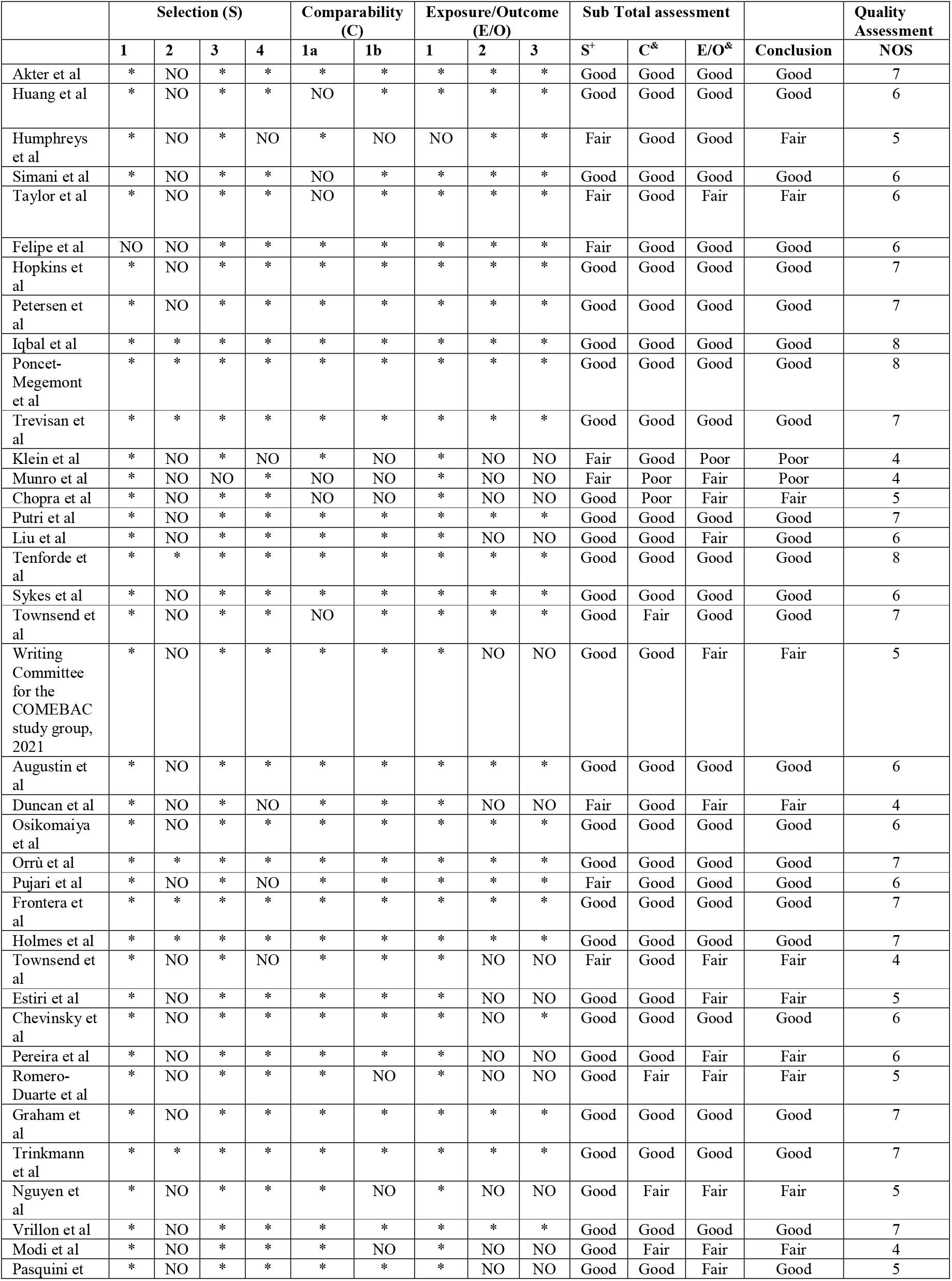
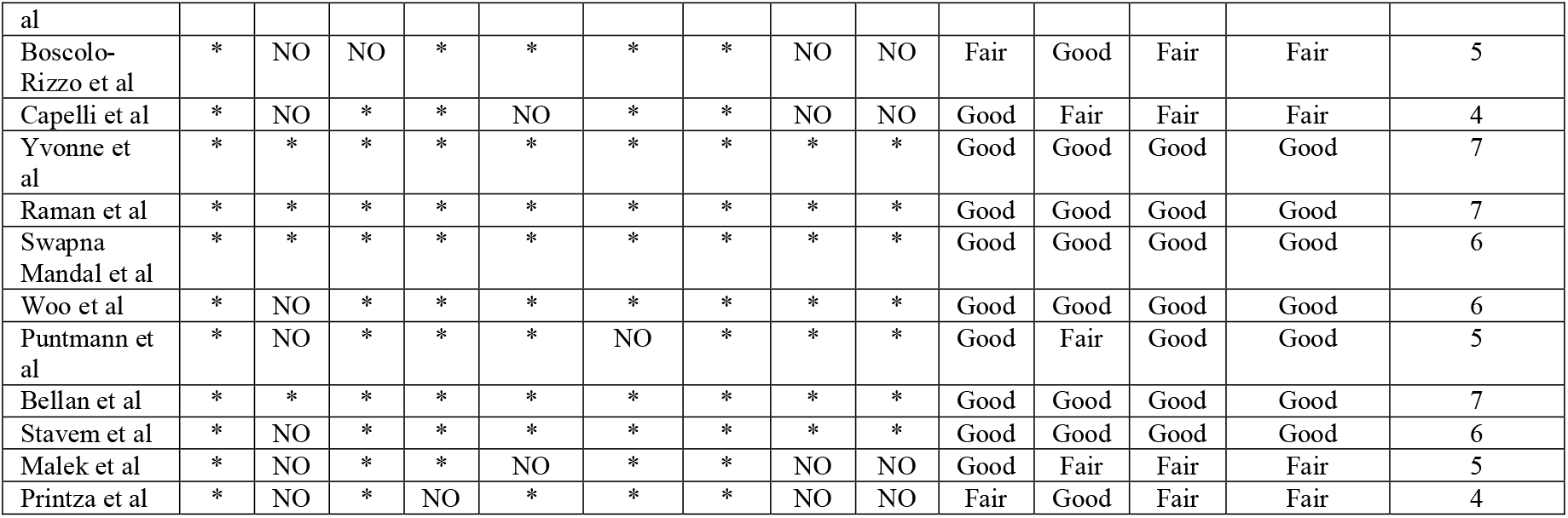
Risk of bias quality assessment

**Table 1b [line 245]:**
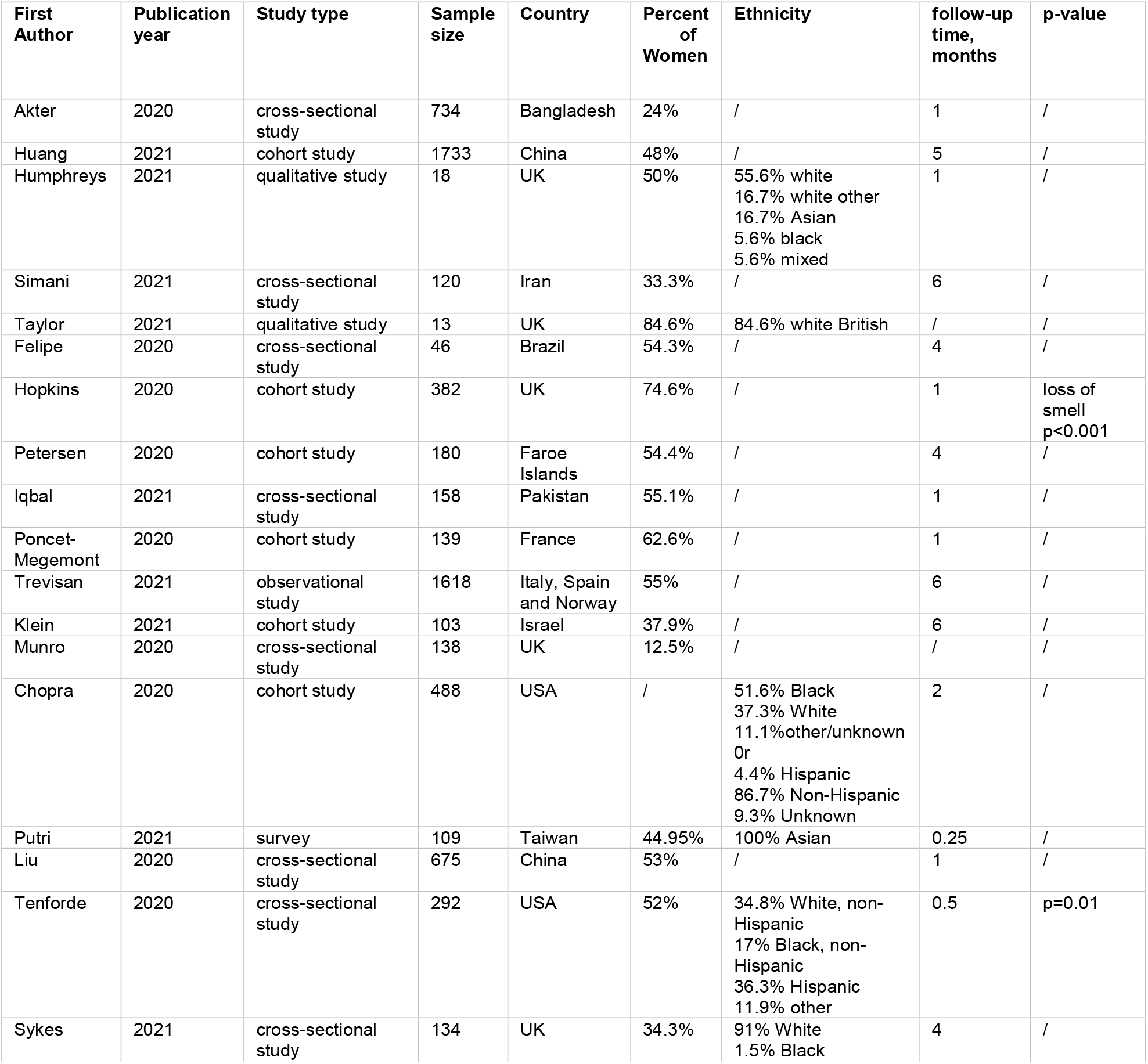
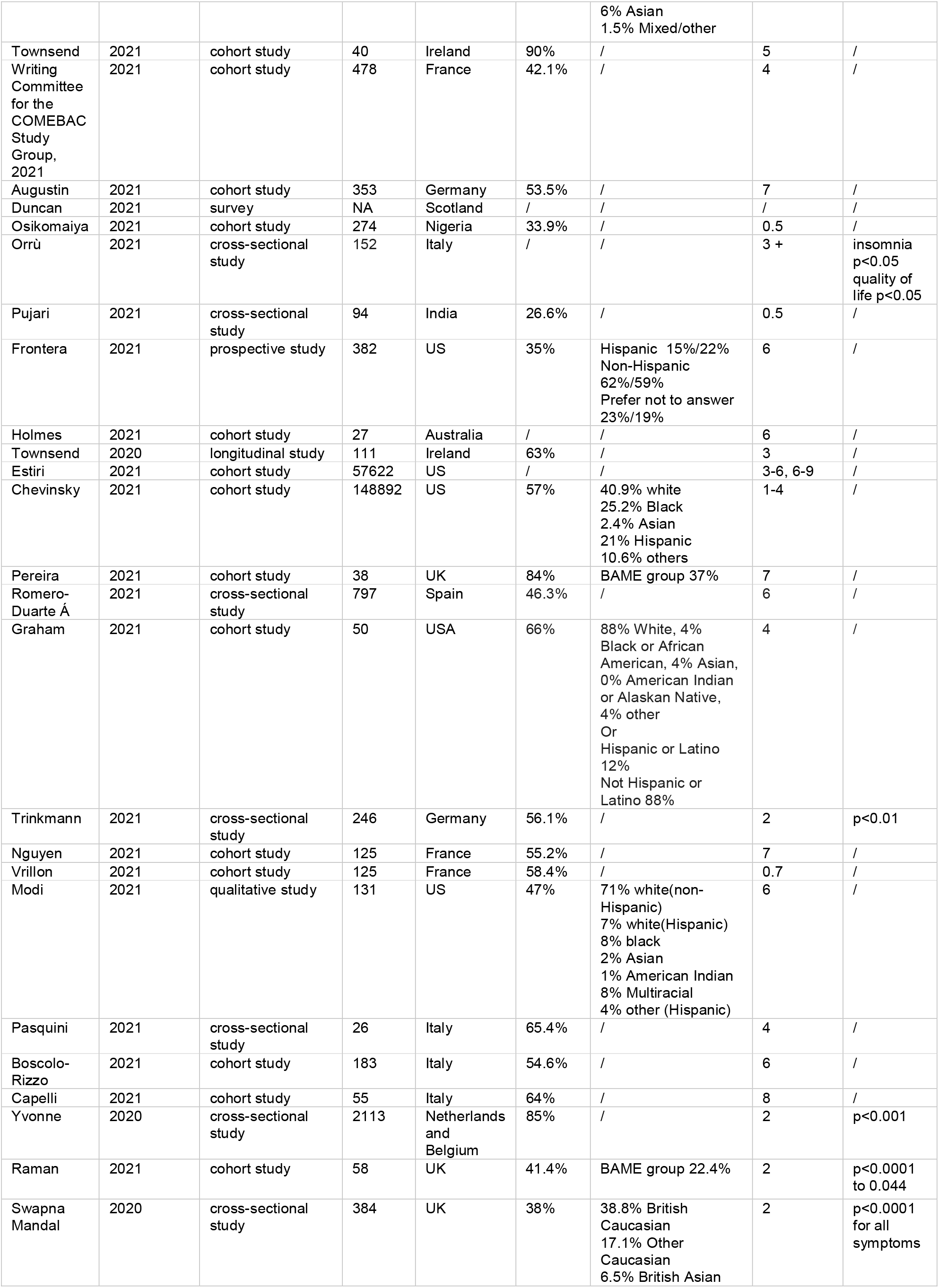
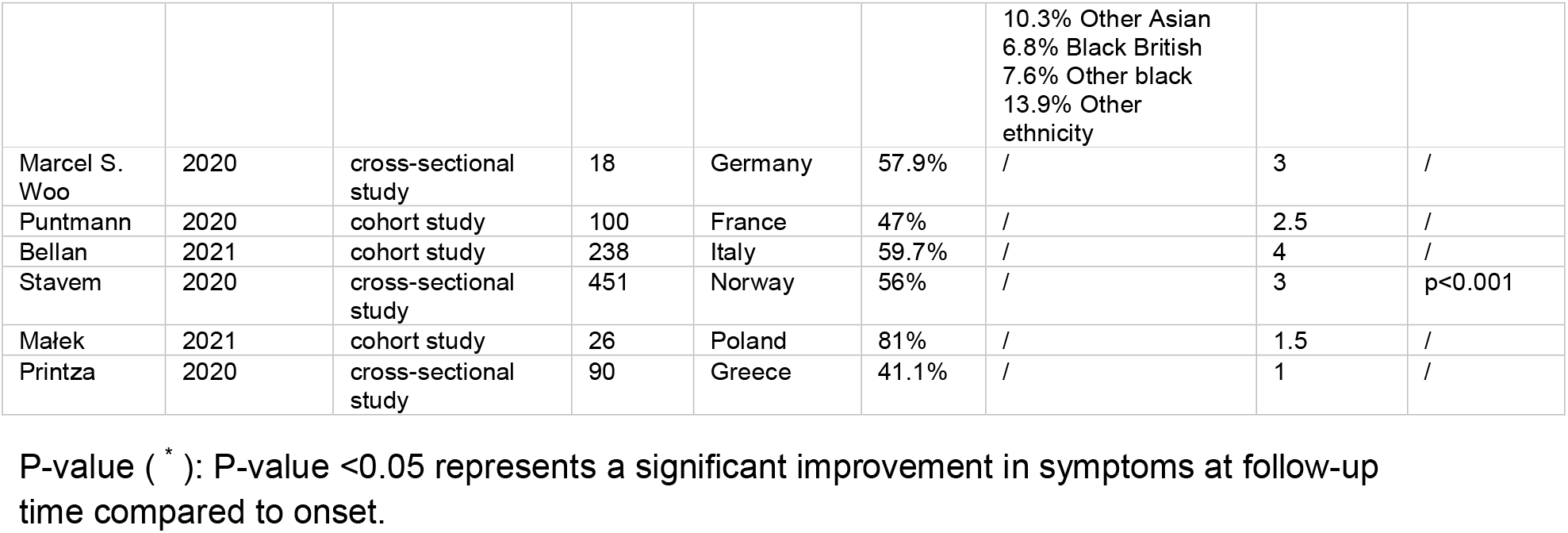
Characteristics of studies included in meta-analysis

- A **Poor** quality score was allocated 0 or 1 star(s) in selection, 0 stars in comparability, and 0 or 1 star(s) in the outcomes domain
- A **Fair** quality score was awarded, 2 stars in selection, 1 or 2 stars in comparability, and 2 or 3 stars in outcomes.
- A **Good** quality score was awarded, 3 or 4 stars in selection, 1 or 2 in comparability, and 2 or 3 stars in outcomes. (6)

### Data analysis

A random-effects model with an inverse variance method was used for the meta-analysis and the heterogeneity was assessed by I^2^. A subgroup analysis was conducted in terms of study geographical location on the symptoms that were reported in more than 10 studies. Sensitivity analysis was used to test the robustness of the results. Funnel plots and Egger’s tests for symptoms with more than 10 studies would be analyzed to detect publication bias. All data analysis will be carried out using R and STATA 15.

## Results

Of the 302 studies identified, 49 met the inclusion criteria. 36 studies were included in the final meta-analysis. This was reported within the PRISMA document as demonstrated in Figure 1.

The 36 studies included comprised of a total sample size of 11,598 people. Of the 36, 50% were cohort studies and the remainder cross-sectional. The longest follow-up time among the 36 studies was 8 months, although the most common follow-up time was 4 months. The 36 studies covered multiple geographical locations, where 19 countries reported five primary classifications of symptomatologies of general clinical, neurological, neuropsychiatry, and cardiopulmonary. Primary clinical features within these categories included fatigue, cognitive impairment, joint pain, anxiety, and depression. These appear to align with the present understanding of LC symptomatologies. Study-based characteristics and outcomes are demonstrated in Table 1b.

### Meta-analysis

The meta-analysis included 36 studies, which are summarized in Figure 2.

**Figure 2:**
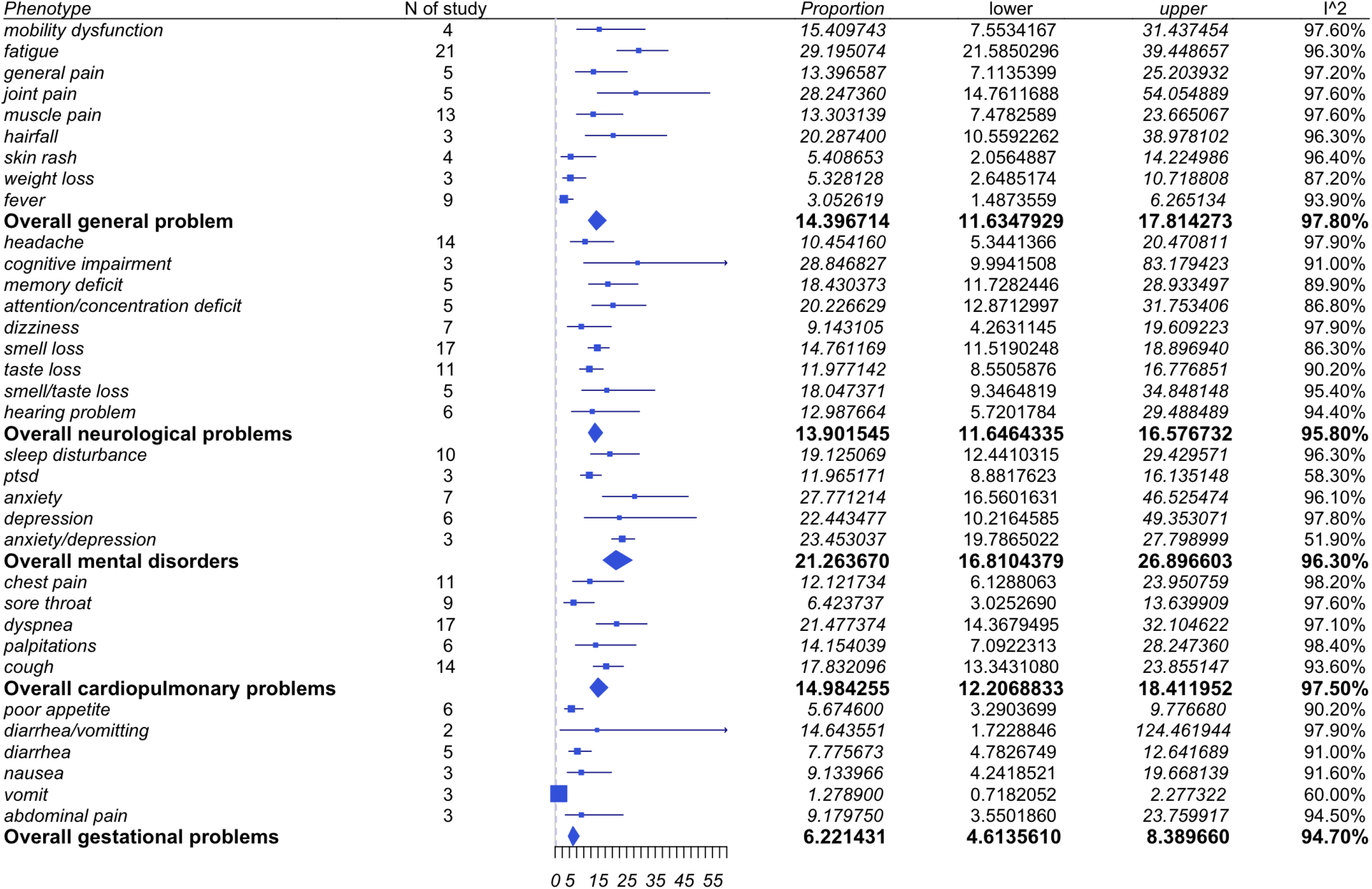
Summary of studies included in meta-analysis.

**Figure 2.1:**
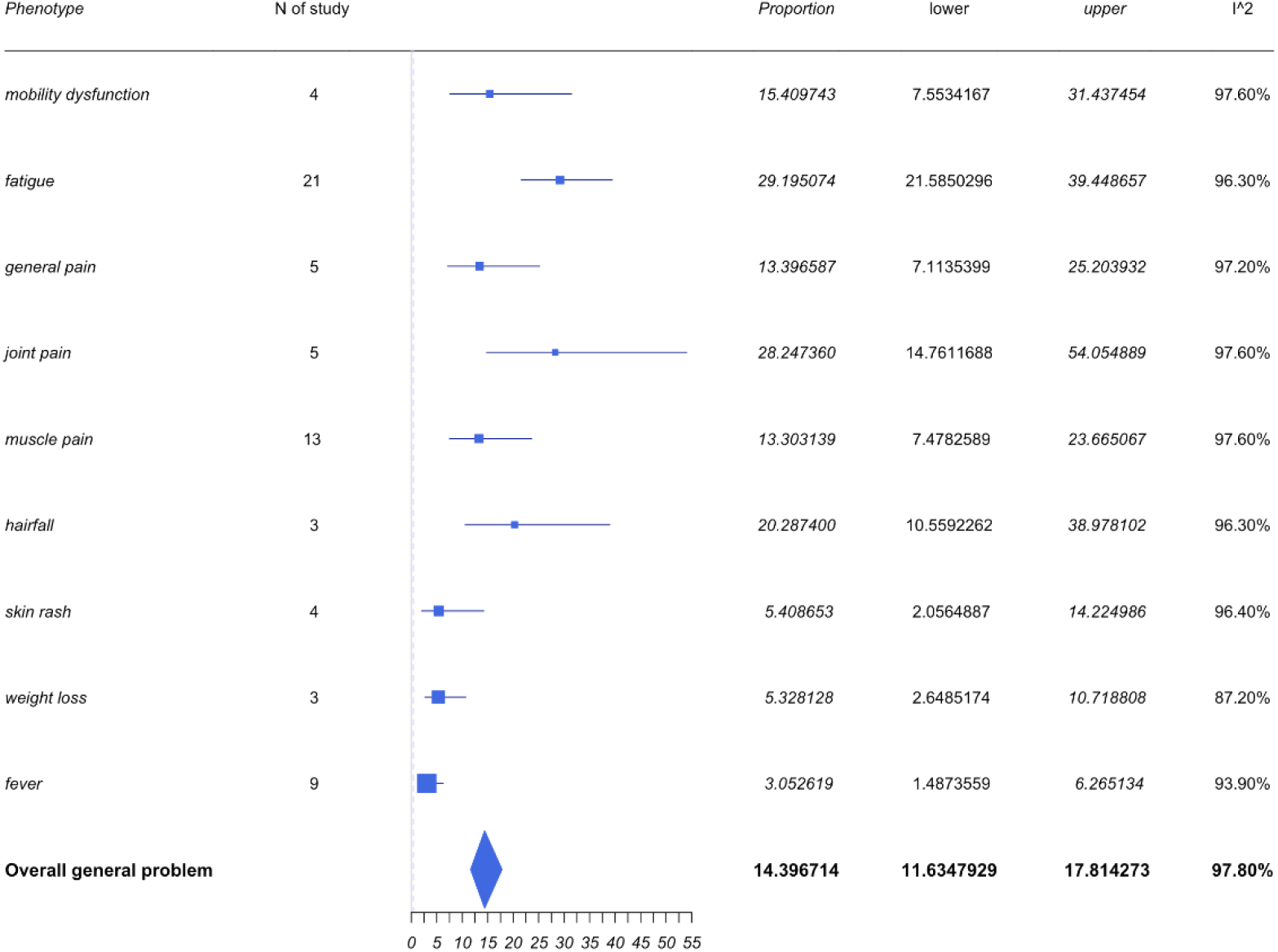
Forest plots for general symptoms.

**Figure 2.2:**
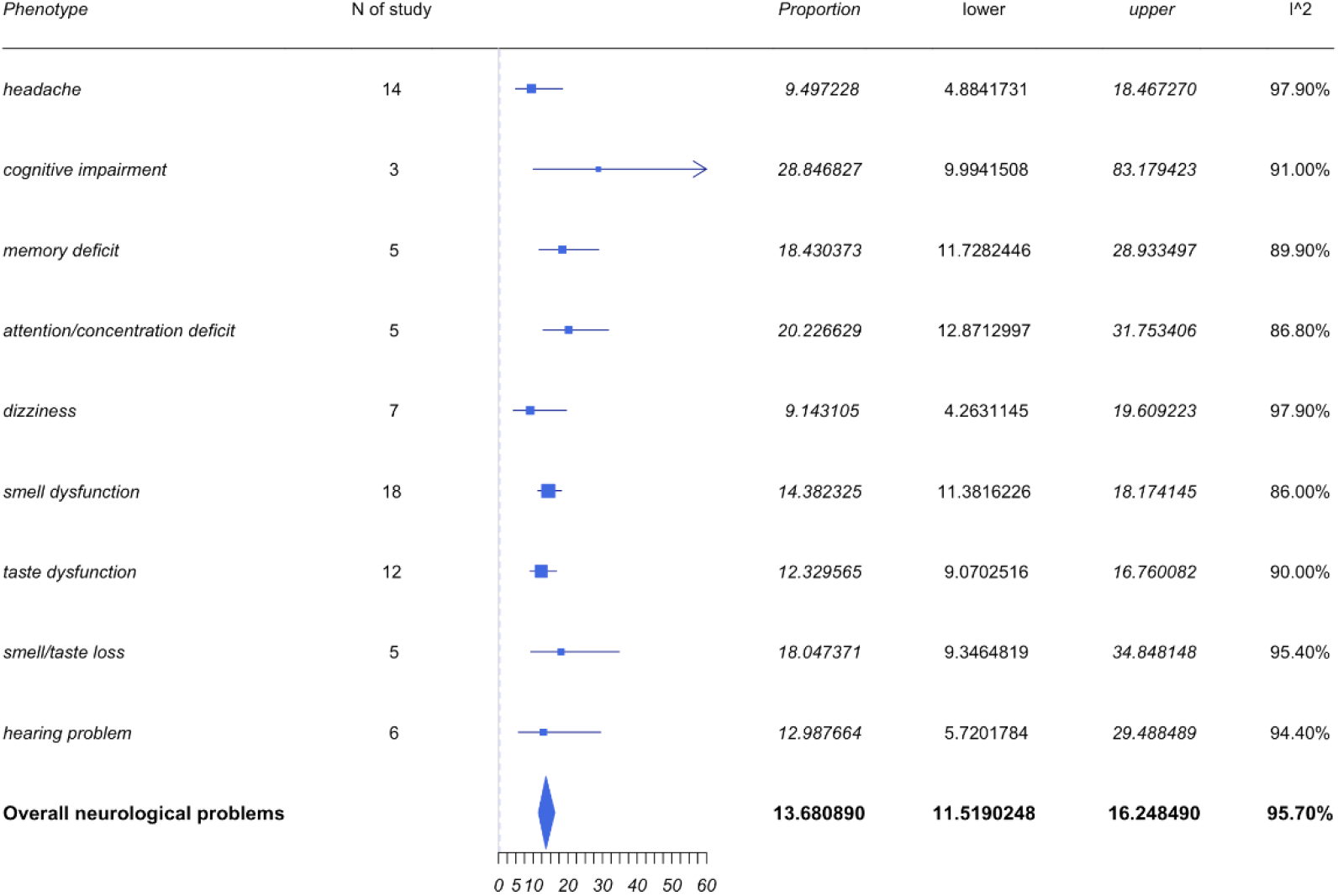
Forest plots for neurological symptoms.

**Figure 2.3:**
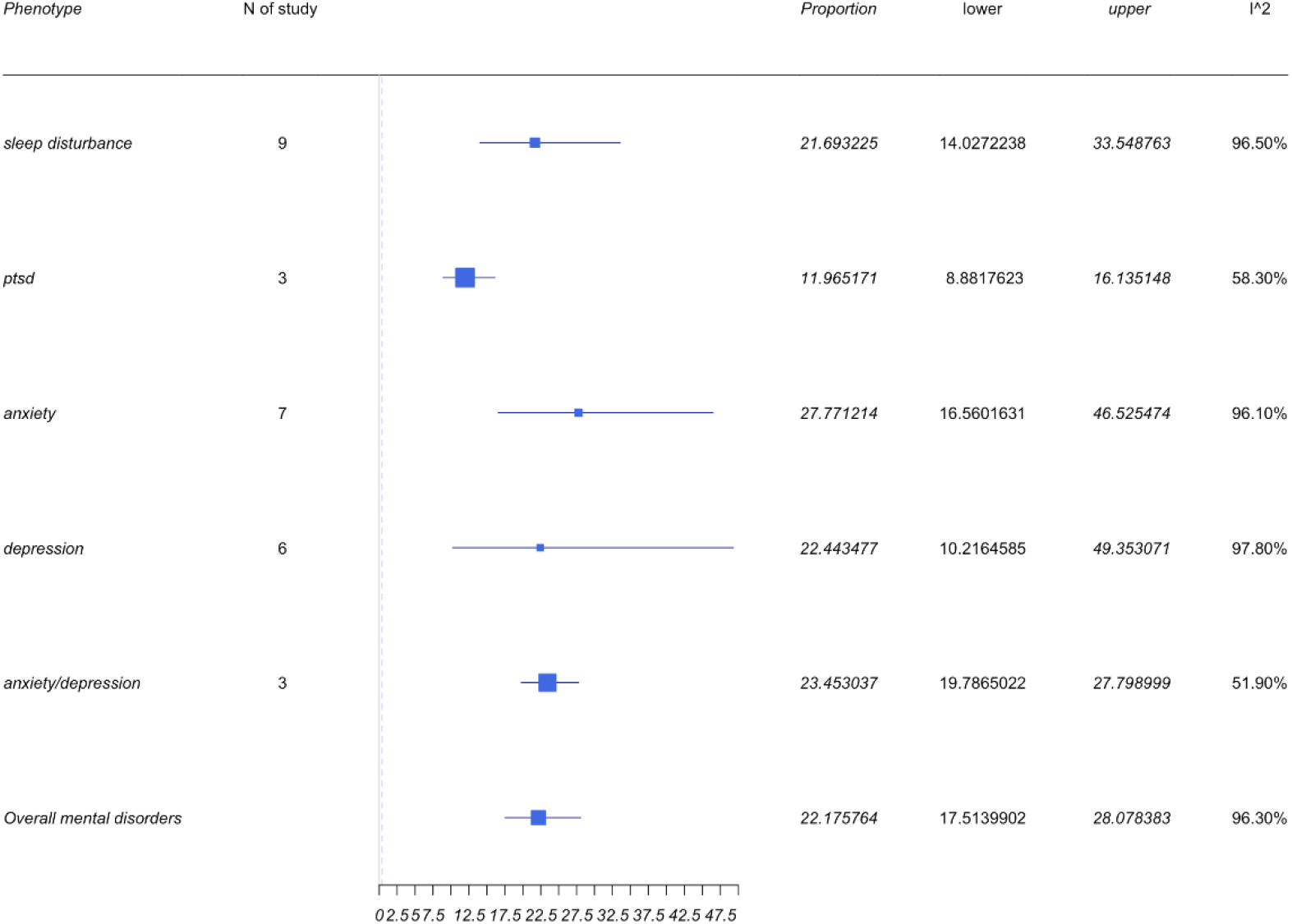
Forest plots for mental health symptoms.

**Figure 2.4:**
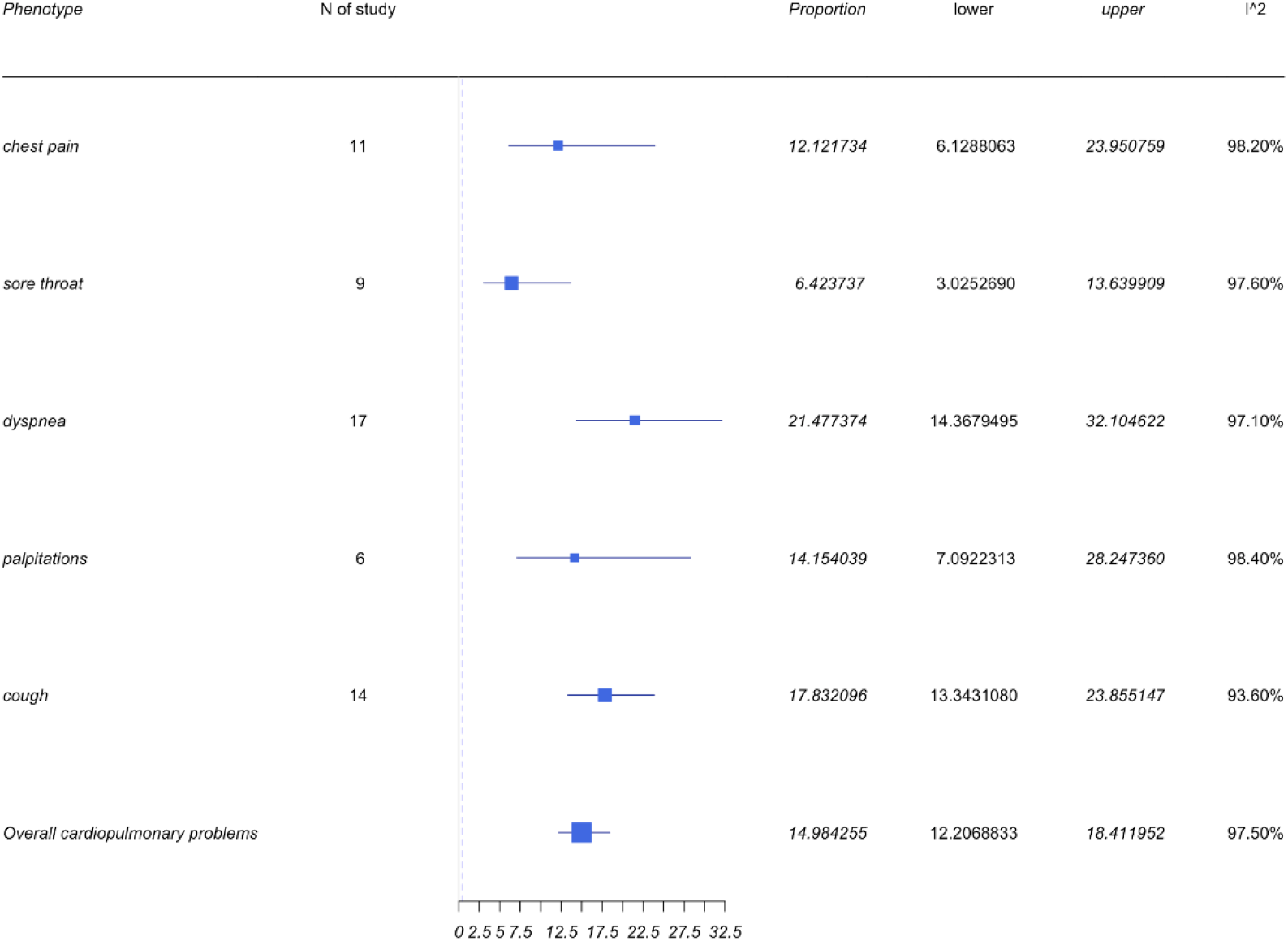
Forest plots for cardiopulmonary symptoms.

**Figure 2.5:**
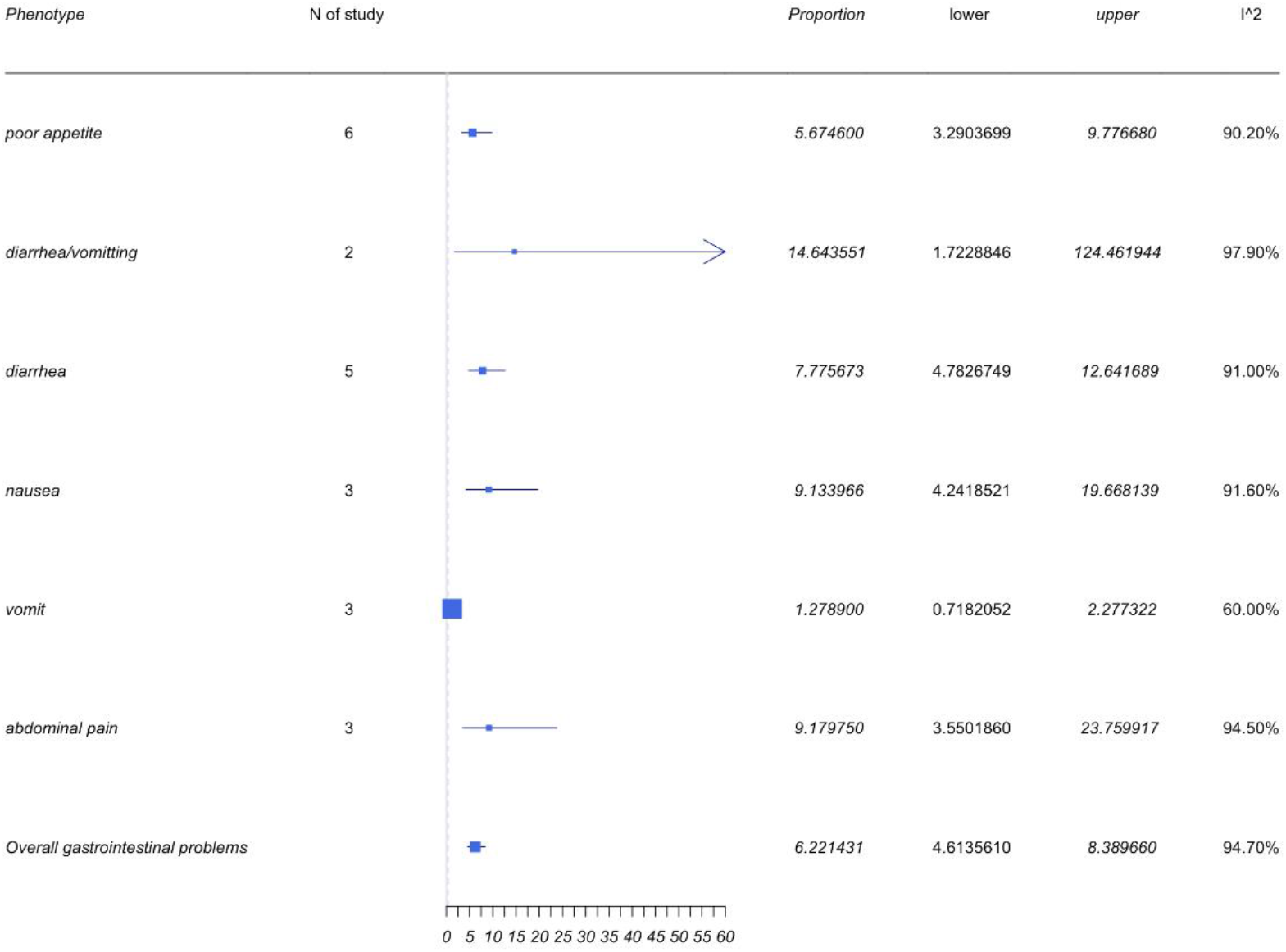
Forest plots for gastrointestinal symptoms.

### Categorization

#### General symptoms

General symptoms included those associated with pain (such as general pain, muscle or joint pain, and mobility dysfunction), fatigue, fever, hair fall, skin rash, and weight loss. The pooled prevalence of the general problem was 14.4% with a 95%CI of 11.63% to 17.81%. A forest plot for general symptoms is shown in Figure 2.1.

Fatigue was the most frequently reported symptom within the general problem category. Twenty-one of the thirty-six studies reported fatigue symptoms and the pooled prevalence of fatigue was 29.2% with a 95%CI of 21.59% to 39.45%. Muscle pain was the second most prevalent symptom reported among the 13 studies, which led to a pooled prevalence of 13.30% with a 95%CI of 7.48% to 23.67%. However, the prevalence rate of muscle pain is not as high as some of the other symptoms associated within the generalized category.

The pooled prevalence of joint pain and hair fall were 28.25% (95%CI 14.76% to 54.05%) and 20.29% (95%CI 10.56% to 38.98%) respectively. It appears that the prevalence of these two symptoms were high, but only a few studies mentioned these in comparison to those reporting fatigue and muscle pain. Therefore, it is worth standardizing these variables across all studies to manage a better understanding of the clinical relevance.

### Neurological symptoms

The neurological symptoms included headache, cognitive impairment, and loss of smell, taste, and hearing. As shown in Figure 2.2, the most frequently reported neurological problems were loss of smell and taste, and headache. The pooled prevalence for loss of smell or taste and both taste and smell as well as headaches were 14.76%, 11.98%, 18.05%, and 10.45% respectively. However, the most prevalent neurological symptom reported appears to be cognitive impairment with a pooled prevalence of 28.85% with a 95%CI of 9.99% to 83.18%. The 95% CI is wide, and the identified heterogeneity based on I^2^ was 91%. Despite the high heterogeneity, only 3 studies mentioned the symptoms of cognitive impairment. Further studies and improved sampling would be required to demonstrate a more precise statistical conclusion in regard to cognitive impairment and LC.

### Mental health symptoms

Four symptoms, and mental health (MH) symptoms including depression, anxiety, PTSD, and sleep disturbances, were reported within the neuropsychiatry category. The pooled results can be found in Figure 2.3. The collective prevalence of MH symptoms was 21.26% (95%CI 16.81% to 26.9%), while each symptom independently also demonstrated a high prevalence. Anxiety prevalence was identified to be 27.77% with a 95%CI of 16.56% to 46.53%, while the prevalence of depression was 22.44% (95%CI 10.22% to 49.35%). The pooled prevalence of studies reporting patients with both anxiety and depression was 23.45% (95%CI 19.79% to 27.8%). The prevalence of sleep disturbance was identified to be 19.13% with a 95%CI of 12.44% to 29.43%. This is an important facet to demonstrate given that there is a large number of studies demonstrating depression and anxiety to be the most commonly reported MH outcomes among SARS-CoV-2 patients.

**Figure 3:**
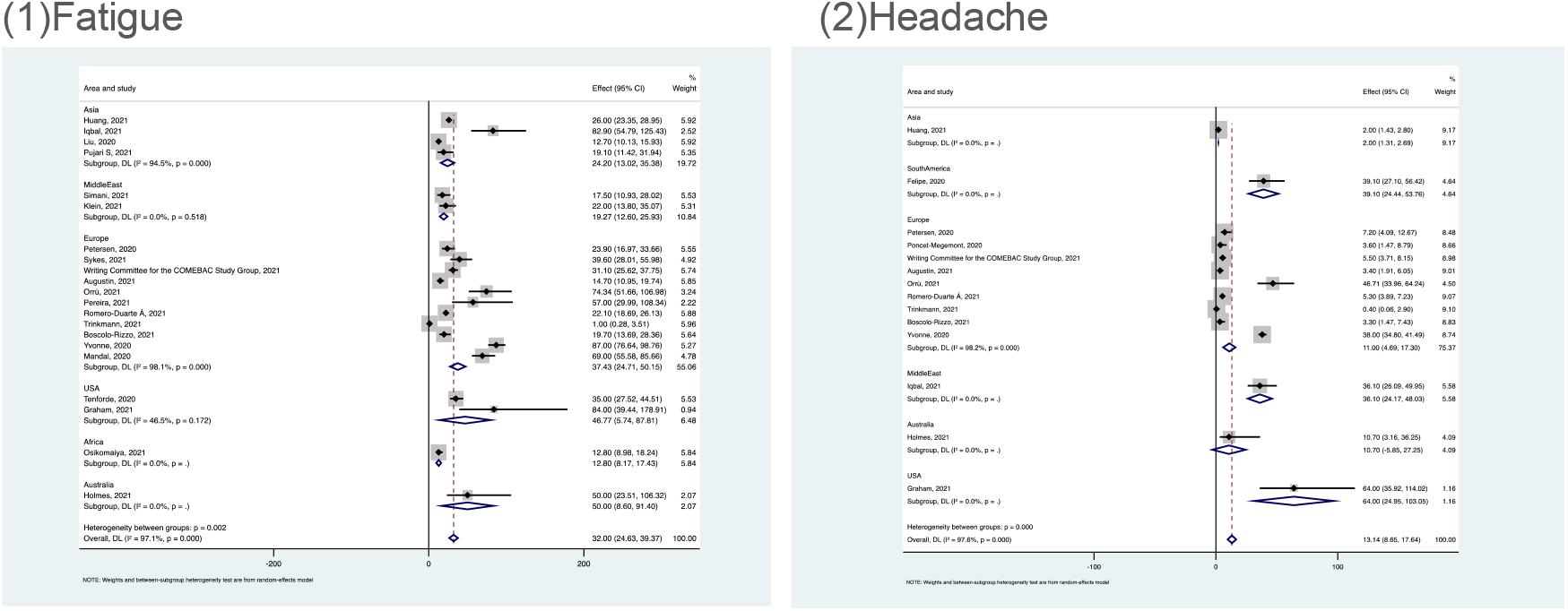
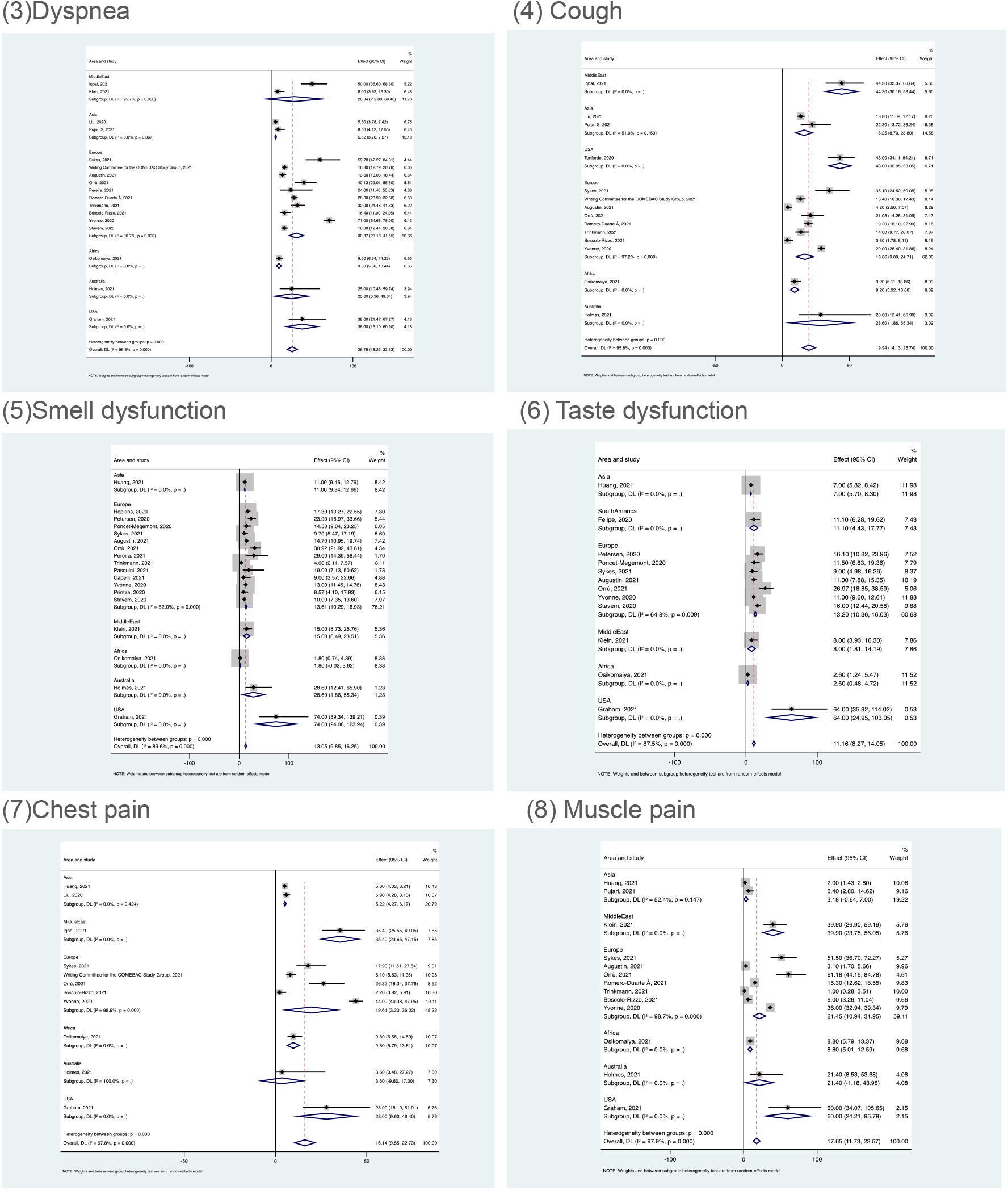
Forest plots of subgroup analysis.

### Cardiopulmonary symptoms

LC patients demonstrated cardiopulmonary symptoms with 5 commonly reported issues of chest pain, sore throat, dyspnea, palpitations, and cough. As can be seen from Figure 2.4, dyspnea appeared to have the highest prevalence with 17 of 36 studies reporting it as a primary end point. The pooled prevalence was therefore 21.48% with a 95%CI of 14.37% to 21.2%. Cough was the second most commonly reported symptom across 14 of 36 studies. The pooled prevalence was 17.83% with a 95%CI of 13.34% to 23.86%.

**Figure 4:**
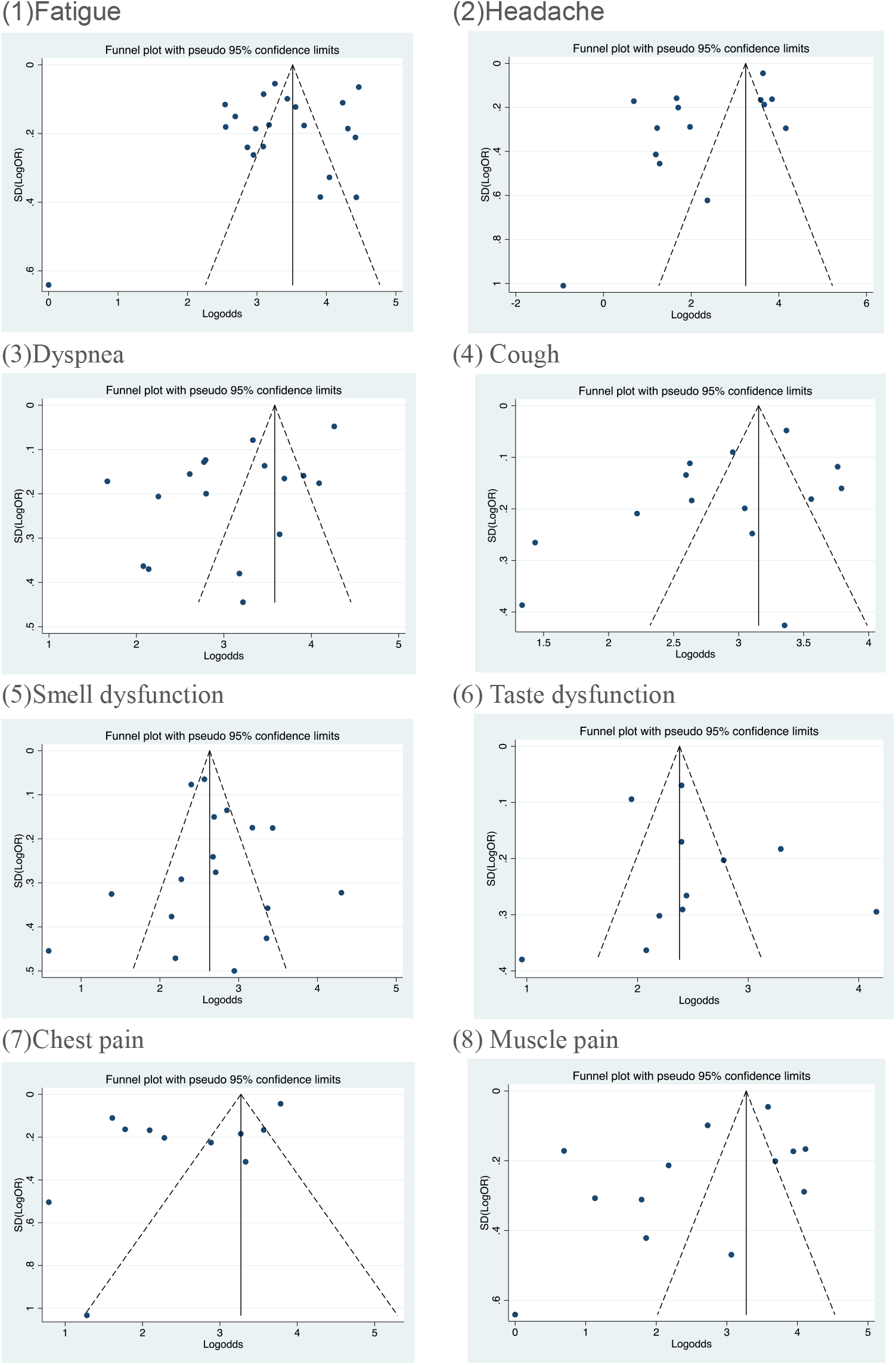
Funnel plots of eight symptoms (reported in more than 10 studies)

### Gastrointestinal symptoms

The overall prevalence of gastrointestinal problems, as shown in Figure 2.5, was 6.22% with a 95%CI of 4.61% to 8.39% and is comparatively minimal to the other categorical symptoms identified. Commonly reported symptoms reported in this category were poor appetite, diarrhea and emesis, diarrhea or emesis, nausea, and abdominal pain. Diarrhea and emesis had the highest prevalence of 14.64% with a 95%CI of 1.72% to 124.46%. Studies about diarrhea/emesis were too small. Only two studies mentioned diarrhea and emesis, which also indicated a high heterogeneity with an I^2^ =97.9%.

**Figure 5:**
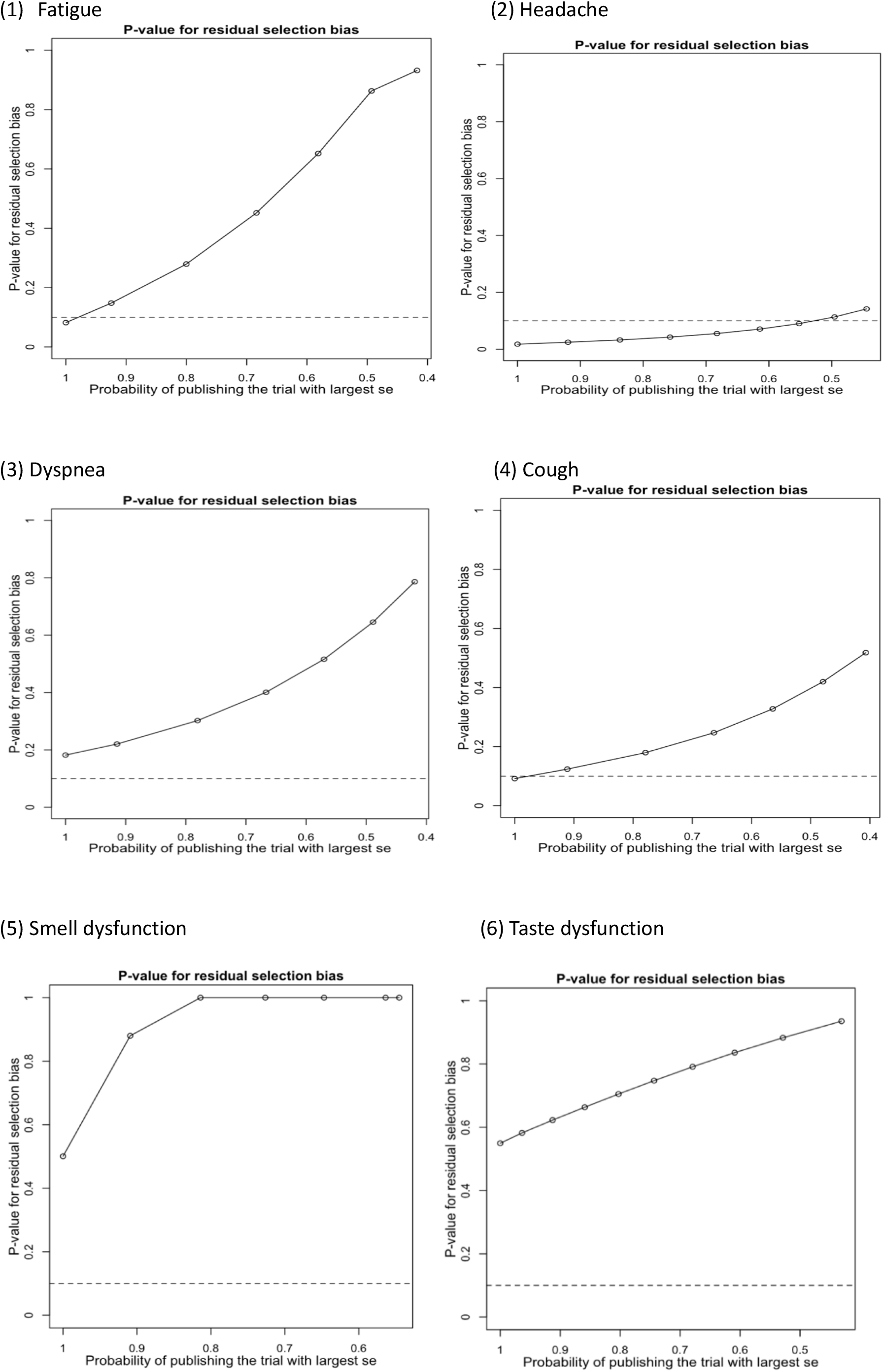
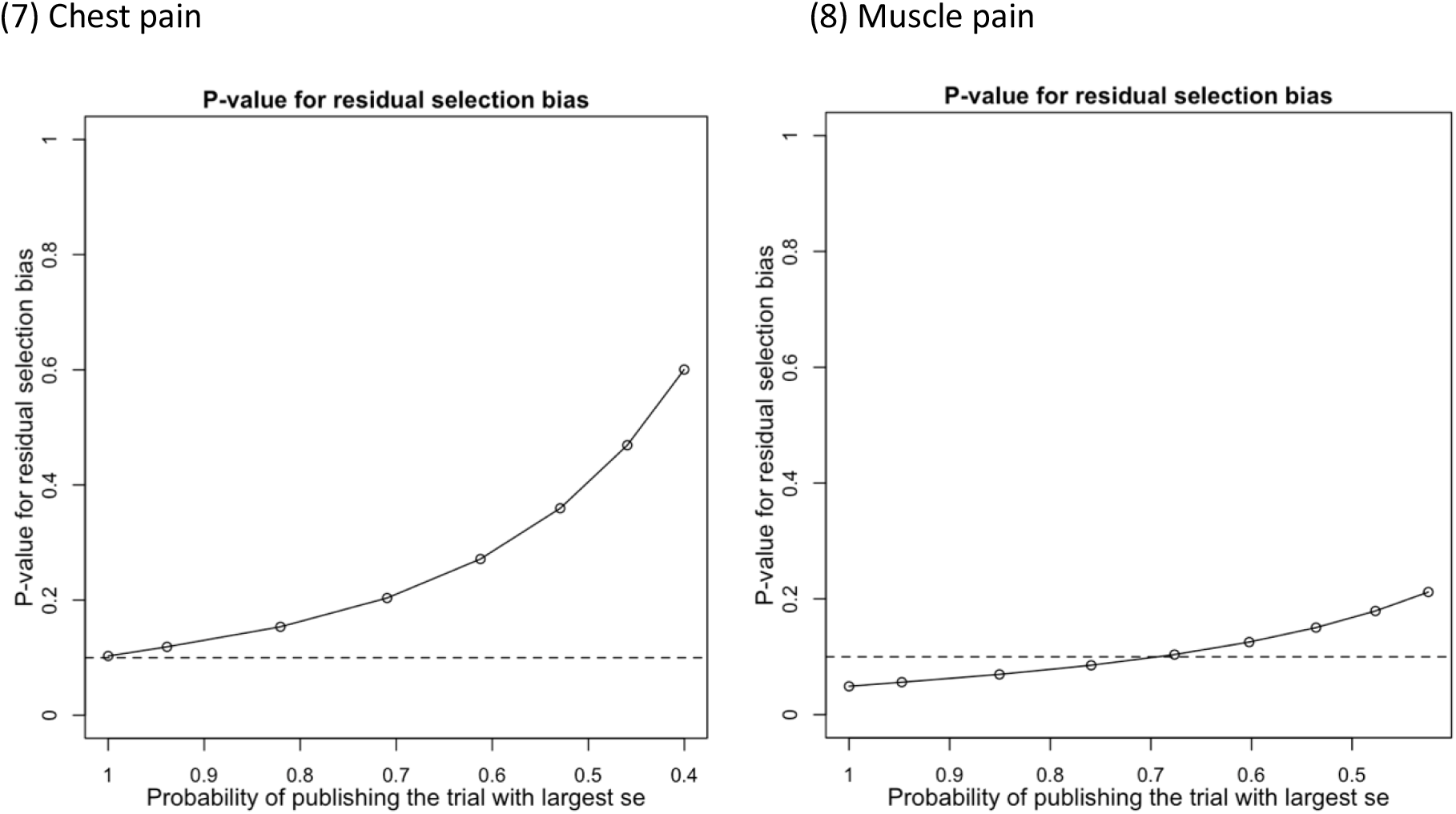
P-values for residual selection bias.

### Subgroup analysis

A subgroup analysis was conducted based on geographical regions correlated with the 8 symptoms of fatigue, headache, cough, loss of smell and taste, dyspnoea, chest, and muscle pain (see Figure 3).

High prevalence of each symptom was reported by the studies from North America (mainly USA), followed by the Middle East and Australia. Most of the symptoms had a lower prevalence in Africa and Asia. Due to the small number of studies in the subgroup, the conclusions may have bias; for this reason, data from one subgroup was of concern to us. 10 studies from Europe reported dyspnoea in this subgroup and the pooled prevalence of this subgroup was 30.87% with 95%CI of 20.18% to 41.55%, which was the second highest prevalence among different regions. This suggested that dyspnoea was a highly prevalent symptom in European countries and should be addressed by the healthcare system to improve post-discharge care.

Funnel plots of the eight symptoms identified were reported in Figure 4. It is apparent, based on the funnel plots, there is the presence of high heterogeneity. Many studies were outside the scope of 95% CI, so it was difficult to intuitively detect the bias. Therefore, Egger’s test was used to determine publication bias.

### Sensitivity analysis

Many studies were outside the 95% confidence interval as demonstrated within the funnel plots, which could impact the overall conclusions of this study. Therefore, a sensitivity analysis was conducted to determine the consensus of the overall conclusion of the study. A Copas selection model was used (8,9) to adjust the pooled prevalence, as demonstrated in Table 2.

**Table 2:** Summarized results of sensitivity analysis

| Phenotype | N of study | Model | Probability of publishing study with largest standard error | Proportion(%) | lower(%) | upper(%) | p-value for differences between two conclusions |
| --- | --- | --- | --- | --- | --- | --- | --- |
| headache | 14 | copas selection model | 49.55% | 13.4637 | 10.0332 | 18.0672 | 0.1135 |
|  |  | random effects model |  | 9.4972 | 4.8842 | 18.4673 |  |
| smell dysfunction | 18 | copas selection model | 100.00% | 14.1682 | 10.1188 | 19.8380 | 0.5006 |
|  |  | random effects model |  | 14.3823 | 11.3816 | 18.1741 |  |
| taste dysfunction | 12 | copas selection model | 100.00% | 12.2338 | 8.2962 | 18.0402 | 0.5495 |
|  |  | random effects model |  | 12.3296 | 9.0703 | 16.7601 |  |
| chest pain | 11 | copas selection model | 100.00% | 12.4236 | 7.2232 | 21.3681 | 0.103 |
|  |  | random effects model |  | 12.1217 | 6.1288 | 23.9508 |  |
| dyspnea | 17 | copas selection model | 100.00% | 21.5591 | 15.1515 | 30.6797 | 0.1819 |
|  |  | random effects model |  | 21.4774 | 14.368 | 32.1046 |  |
| cough | 14 | copas selection model | 91.11% | 18.176 | 12.8109 | 25.7852 | 0.1238 |
|  |  | random effects model |  | 17.8321 | 13.3431 | 23.8551 |  |
| fatigue | 21 | copas selection model | 92.46% | 29.1951 | 21.5850 | 39.4487 | 0.1478 |
|  |  | random effects model |  | 28.9161 | 20.3158 | 41.1531 |  |
| muscle pain | 13 | copas selection model | 67.65% | 15.6426 | 9.1963 | 26.6103 | 0.1038 |
|  |  | random effects model |  | 13.3031 | 7.4783 | 23.6651 |  |

In Table 2, the proportion of selected studies varied, and the changes in the P-value of the residual selection bias are depicted in Figures 5 (1)-(8). The Copas model (CSM) was used to determine bias within studies based on P-values exceeding 0.1. The proportion of studies used within the CSM are listed within Table 2. It is evident the CSM selected 49.55% studies with headache as a symptom, while the remaining 50.45% indicated a significant standard error, demonstrating poor quality and high heterogeneity, thus were excluded.

The result from the CSM was compared to a random effects model (REM), indicating P-values exceeding 0.05, which demonstrates a lack of statistical significance. Therefore, the results of this study are consistent and provide robust conclusions.

### Publication bias

Egger’s test was used to determine publication bias. The P-values were calculated based on Egger’s test.

As shown in Table 3, studies reporting symptoms of headache and dyspnea have significant bias, with P-values of 0.022 and 0.007 respectively. Therefore, the pooled prevalence of headache and dyspnea were 9.5% and 21.48%. In Figures 4(2) and (3), the prevalence of headache and dyspnea may have been underestimated, and more studies should be found to further confirm the conclusions.

**Table 3:** Summarized P-values of Egger’s tests for symptoms with more than 10 studies

| Phenotype | Number of studies | P-value of Egger's test |
| --- | --- | --- |
| **General problems** |  |  |
| fatigue | 21 | 0.14 |
| muscle pain | 13 | 0.093 |
| **Neurological problems** |  |  |
| Smell loss | 18 | 0.616 |
| Taste loss | 12 | 0.517 |
| headache | 14 | **0.022** |
| **Cardiopulmonary problems** |  |  |
| Chest pain | 11 | 0.048 |
| dyspnea | 17 | **0.007\*** |
| cough | 14 | 0.085 |

Note: ( \* ) : p<0.05 indicates significance

## Limitations

There are strengths and weaknesses to our study given that comparing patients with maximum symptoms risks bias reporting. Studies that reported on neuropsychiatry symptoms of depression and anxiety, for example, did not demonstrate a clinical diagnosis. The identified and reported features cannot be deemed to be LC as these patients could have underlying conditions that may not have been reported. Patients who may have had critical respiratory illness, for example, may have been part of these studies, but this data was not captured within the original peer-reviewed publications. This would influence the analysis conducted within our study; therefore, an underrepresentation and/or overrepresentation of some of these symptoms is a point to consider.

## Discussion

This meta-analysis demonstrates the most recent studies identified with possible long COVID symptoms. The pooled data indicate both self-reported and clinically reported symptoms. This initial step is vital to design and develop comprehensive research in the future, especially since SARS-CoV-2 appears to be reporting a varying degree of symptoms.

The evidence identified demonstrates that long COVID appears to have multiple symptoms, without clear aetiology similar to fibromyalgia and chronic fatigue syndrome. The forementioned conditions also have an association with postviral illness which appear to last longer than previously anticipated. As a result, healthcare systems endeavour challenges with draining resources and souring costs in addition to wellbeing concerns for staff. Another direct result of long COVID disease will be the added burden on waiting times for patients requiring other clinical care and elective procedures created by the pandemic.

The population prevalence of long COVID identified here could be used to determine symptom-based models to evaluate the requirement for healthcare system resources, and possible disease sequalae which may require care. Presently, instituting effective therapies is based upon present clinical knowledge than evidence-based practices. Repurposing drugs is another common theme among clinicians, and these raise concerns around long COVID potentially becoming a chronic condition in the near future, especially for patients who had significant issues with COVID. With a growing number of variants of SARS-Cov-2 virus, this further exacerbates the present unknowns of managing these patients in an optimal manner. However, this meta-analysis does provide an opportunity to plan early intervention strategies and target therapies in the future.

As it is an evolving pathology, further studies are being reported and published swiftly, which has its own challenges. Therefore, to consistently report the latest evidence, there is a requirement for a *living systematic review and meta-analysis* as well as better methodologies should be developed. It is interesting to note that developed countries appear to have a higher incidence of long COVID based on the geographical data identified within this study. There is a possibility of over- and under reporting, as well as validation of self-reported data. The lack of accurate validated measurements for reported long COVID symptoms similar to other fundamental clinical measures such as blood pressure and temperature causes further problems.

In addition to these factors, ethical and moral implications to patients, the public, and healthcare professionals continue to augment debates as the pandemic has forced all stakeholders to rethink access to healthcare and treatment.

It is evident from this study that there are post-COVID symptoms that patients continue to report. It might be beneficial to reduce the severity of the disease. A useful method to reduce these of course would be to increase the vaccination program outputs globally. With mass migration also attributed to the spread of Covid-19, an important facet to consider would be to understand the barriers and potential issues around vaccine acceptability, especially for those returning to work or their education in countries of residence.

The COVAX Facility is an international collaborative effort shared between the Coalition for Epidemic Preparedness Innovations (CEPI), the Global Alliance for Vaccines and immunizations (GAVI), the World Health Organization, supporting governments and international organizations. (10) The COVAX Facility is meant to facilitate the development and production of diagnostics, therapeutics, and vaccines to combat the COVID-19 pandemic and to make them accessible to LMIC governments. (11,12) The COVAX Facility does not have a legal entity, therefore cannot enter into binding agreements, and relies upon agreements between its constituent partners (e.g. GAVI, WHO) procuring government, and the vaccine manufacturers. (13) Therefore, the law of contract governs access to vaccines, data related to vaccine procurement and distribution, and related matters.

Similarly, sharing of data to better assess the mental and physical health sequalae has been hampered by the lack of an international coordinating mechanism to do so or a uniform set of guidelines that governments, public health officials, private companies, and others may use to share such data. (14) As a result, COVID data related to incidence, disease burden, and long COVID as well as a potential disease sequalae may not be fully understood by the global healthcare community. This is a particular a problem for assessing both COVID and long COVID syndrome impact on differing ethnicities, age groups, and overall health status. Even within academic and clinical research, only open access publications provide insight into evidence.

The justification of resources being reallocated to non-life-threatening sequelae of long COVID could be a contentious topic. This further raises legal implications for policymakers.

The research on nociplastic and immunological explanations for pain symptoms could throw more light in future on the development of pain with long COVID. Genetic studies may also throw some light into the development of long-standing chronic pain or even long COVID, although this requires bio-sampling at a high frequency. The role of nutritional status and activity levels also needs to be established and its association with long COVID needs further study.

## Conclusions

A key finding of this study is that the speed at which SARS-CoV-2 research is being conducted has meant epistemic authority consolidates around particular clinical areas. Therefore, it is vital to synthesise the evidence without any background noise. However, as demonstrated in this study, the gathering of LC data has been limited. The identified data could be associated with autonomic dysfunction, although to confirm this, further investigations would be required. Mapping LC outcomes would be a long-term commitment; therefore, future systematic reviews and meta-analysis should be reported in a *living* format, combining both clinical and research data to allow a more comprehensive synthesis of evidence with a view to using surveillance data.

## Data Availability

All data produced in the present study are available upon reasonable request to the authors

## List of abbreviations

CI: confidence interval
CSM: Copas selection model
GAVI: Global Alliance for Vaccines and Immunizations
LC: Long-Covid
MH: Mental health
NOS: Newcastle-Ottawa Scale
PRISMA: Preferred Reporting Items for Systematic Reviews and Meta-Analysis
REM: Random effects model
WHO: World Health Organization

## Declarations

### Ethics approval and consent to participate

Not applicable

### Consent for publication

Not applicable

### Availability of data and materials

All data used within this study have been publicly available. The authors will consider sharing the dataset gathered upon request.

### Competing interests

PP has received a research grant from Novo Nordisk, and the other, educational from Queen Mary University of London, from John Wiley & Sons, and other from Otsuka, outside the submitted work. SR reports research funding associated with other studies from Janssen, Otsuka, and Lundbeck. AS reports funding associated with other projects from Medtronic. GD reports research funding associated with NIHR RCF. VR reports funding associated with other studies from the Medical Research Council.

This research is based on evidence gathered systematically and has not been influenced unduly by expertise.

All other authors report no conflict of interest.

The views expressed are those of the authors and not necessarily those of the NHS, the National Institute for Health Research, Department of Health and Social Care or Academic institutions.

### Authors’ contributions

PP and GD developed study design and GD wrote the first draft of the manuscript. AN conducted database searches and study selection and data extraction. YZ, JQS, GD performed statistical analyses and contributed to the results’ section. AN, AS, GD, YE, YZ, DK, VR, SR, SH, KE, JQS and PP critically reviewed and revised the manuscript. All authors approved the final version of the manuscript.

## Acknowledgements

The authors acknowledge initial contributions made by Dr M Sam Chong, Dr Robert Shane Delamont and Dr Mayur Bodani. We would like to acknowledge Professor Balakrishna Shetty for providing us his thoughts on Covid and Long-covid as a practicing physician managing the ongoing care of patients.

This paper is part of the multifaceted EPIC project, sponsored by Southern Health NHS Foundation Trust and in collaboration with the University of Oxford, University College London, University College London NHS Foundation Trust and Southern University of Science and Technology (China).

